# Direct therapeutic effect of sulfadoxine-pyrimethamine on nutritional deficiency-induced enteric dysfunction in a human intestine chip

**DOI:** 10.1101/2023.07.13.23292620

**Authors:** Seongmin Kim, Arash Naziripour, Pranav Prabhala, Viktor Horváth, Abidemi Junaid, David T. Breault, Girija Goyal, Donald E. Ingber

## Abstract

Sulfadoxine-pyrimethamine (SP) antimalarial therapy has been suggested to improve the birth weight of infants in pregnant women in sub-Saharan Africa independently of malarial infection. Here, we investigated whether SP could have a direct impact on improving intestinal function, thereby enhancing the absorption of essential fats and nutrients crucial for fetal growth.

Using a human organ-on-a-chip model, we replicated the adult female intestine with patient organoid-derived duodenal epithelial cells interfaced with human intestinal endothelial cells. Nutrient-deficient (ND) medium was perfused to simulate malnutrition, resulting in the appearance of enteric dysfunction indicators such as villus blunting, reduced mucus production, impaired nutrient absorption, and increased inflammatory cytokine secretion. SP was administered to these chips in the presence or absence of human peripheral blood mononuclear cells (PBMCs).

Treatment with SP successfully reversed many abnormalities observed in malnourished chips, as confirmed by transcriptomic and proteomic analysis. Notably, SP significantly enhanced intestinal absorptive functions. Furthermore, SP suppressed the recruitment of PBMCs in the nutrient deficient chips. SP may improve the birth weight of children born to malnourished mothers by countering the effects of enteric dysfunction and suppressing inflammation. These findings highlight the possibility of using SP as a direct intervention to improve maternal absorption and subsequently contribute to healthier fetal growth.

## INTRODUCTION

Malnutrition during pregnancy, particularly a lack of vital nutrients like iron, fat, calcium, and zinc, can lead to adverse birth outcomes such as low birth weight and developmental delays in children.^1,2^ Low birth weight is also an important indicator of infant mortality, which can be caused by preterm birth, small size for gestational age, and malaria.^2^ To combat malaria infection and reduce the risk of adverse birth outcomes, the World Health Organization (WHO) recommends intermittent preventive treatment with combined sulfadoxine and pyrimethamine (SP) during pregnancy.^3^ Interestingly, a recent study found that maternal SP treatment can reduce the incidence of low birth weight infants even in the absence of malarial infection and when the organism is resistant to SP treatment.^4^ The physiological relevance of this observation remains unclear as there is limited understanding of the effects of SP treatment on maternal health and birth outcomes.

In this study, we aimed to investigate the effects of SP treatment on nutrient absorption in the adult female small intestine and its impact on intestinal function using functional, transcriptomic, and metabolic assays. To achieve this, we employed a microfluidic human organ-on-a-chip model of the Intestine (Intestine Chip) that is lined by human intestinal epithelium derived from organoids isolated from the duodenum of young adult females interfaced with primary human intestinal microvascular endothelium. This model allowed us to directly assess the potential therapeutic effects of SP on the intestine. To simulate malnutrition in the adult female intestine, we exposed the Intestine Chip to nutritionally deficient (ND) medium lacking niacinamide and tryptophan. We have previously demonstrated that human Intestine Chips cultured under these ND conditions exhibited characteristic signs of environmental enteric dysfunction (EED), such as villus blunting, decreased mucus production, and reduced absorption of fatty acids and peptides, as well as altered transcriptomic profiles, consistent with previous clinical findings in EED patients.^5^ Given the finding that SP seems to increase birth weight in maternal patients who are resistant to the anti-malarial effects of this drug,^4^ we administered SP through the apical epithelium-lined channel of the Intestine Chip to mimic oral administration in patients and observed its effects under both healthy and ND conditions. Recognizing the complex interplay between intestinal epithelial cells and the mucosal immune system,^6,7^ we also conducted experiments using human peripheral blood mononuclear cells (PBMCs) introduced into the basal endothelium-lined channel of the Intestine Chips. Our results demonstrate that SP has direct protective effects on the malnourished adult female intestine.

## RESULTS

### SP mitigates the effects of nutritional deficiency in the human Intestine Chip

In this study, we set out to address the adverse effects of nutritional deficiency (ND) using an Intestine Chip model that faithfully replicates the structure and functionality of the human intestine (figure 1A).^9^ Primary duodenal epithelial cells isolated from patient-derived organoids were cultured on the upper extracellular matrix-coated surface of the porous membrane that separates two parallel microchannels within the chips and primary human intestinal microvascular endothelium was grown on the lower surface of the same membrane. The culture medium was continuously perfused through both channels while peristalsis-like deformations were produced by the application of cyclic suction to side chambers within the flexible polymer chip. Recreation of the intestinal microenvironment in this manner leads to the spontaneous formation of intestinal villi-like structures (figure 1B,C, and supplementary figure 1A). The chip also maintained an intact intestinal barrier (figure 1D) and actively produced mucus (figure 1E,F), including Mucin 5B (MUC5B) (figure 1G and supplementary figure 1B).

**Figure 1:**
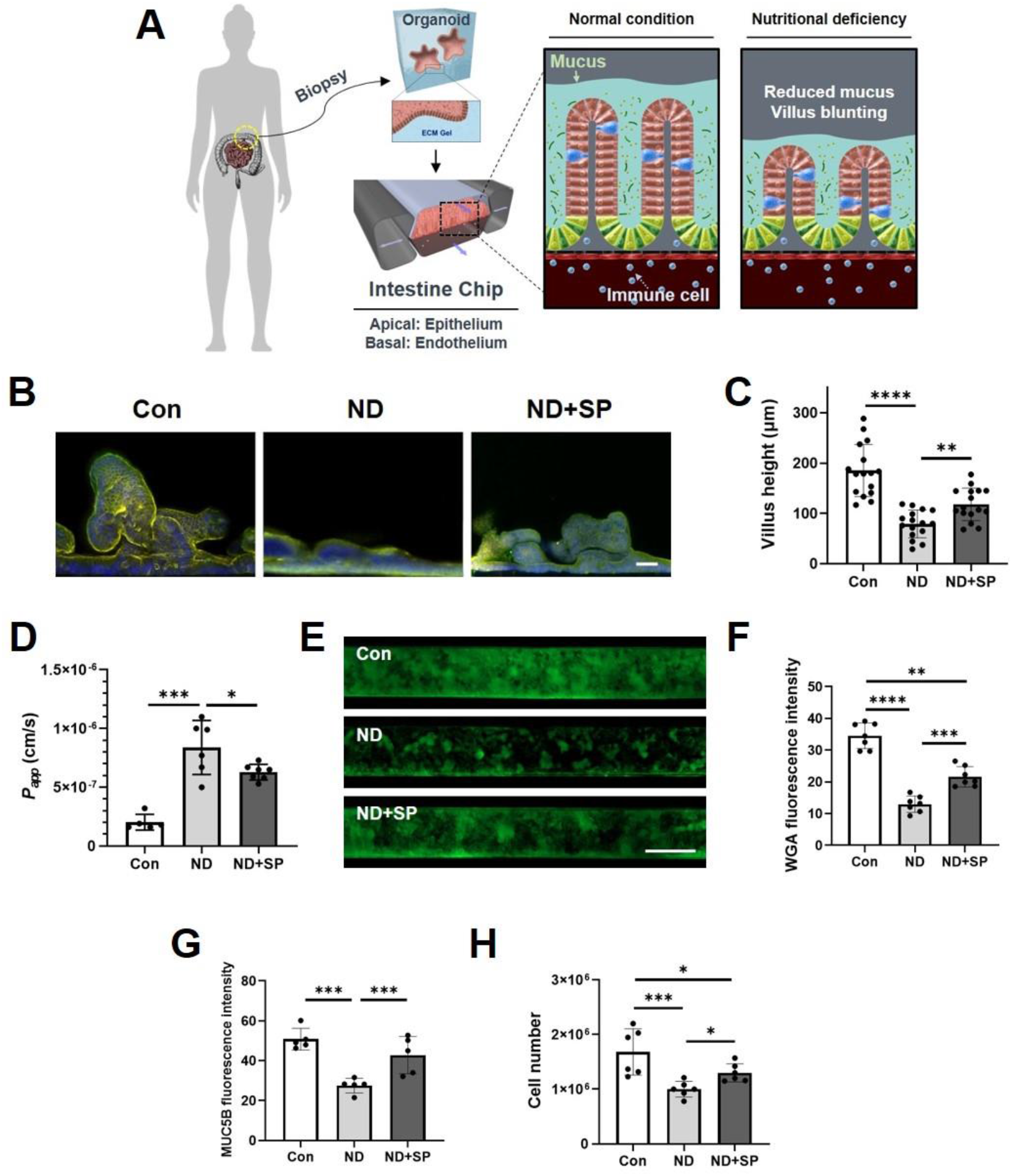
Effect of nutritional deficiency and SP treatment on adult female Intestine Chip. (A) A schematic cross-sectional view of the duodenal organoid-derived Intestine Chip showing the apical (epithelium) and basal (endothelium) microchannels. The sulfadoxine-pyrimethamine (SP) drug combination was applied to the apical channel and the human peripheral blood mononuclear cells (PBMCs) were applied to the basal channel. (B) Immunofluorescence imaging of cross-sectioned Intestine Chips showing villus-like structures, green: phalloidin, yellow: ZO-1, blue: Hoechst 33342. Scale bar = 50 µm. (C) Differences in villus-like structure height between normal (Con), nutritionally deficient (ND), and SP-treated ND (ND+SP) Intestine Chips. \*\**p* < 0·01, \*\*\*\**p* < 0·0001 by Two-tailed Student’s t-test. (D) Apparent permeability (*P_app_*) after SP treatment. \**p* < 0·05, \*\*\**p* < 0·001 by Two-tailed Student’s t-test. (E) Immunofluorescence imaging of Intestine Chips stained with wheat germ agglutinin (WGA) with Alexa-488 conjugated lectin showing mucus production. Scale bar = 200 µm. (F) Differences in fluorescence intensity of WGA. \*\**p* < 0·01, \*\*\**p* < 0·001, \*\*\*\**p* < 0·0001, by Two-tailed Student’s t-test. (G) Differences in fluorescence intensity of MUC5B. \*\*\**p* < 0·001 by Two-tailed Student’s t-test. (H) Comparison of cell number between Con, ND, and ND+SP Intestine Chips. \**p* < 0·05, \*\*\**p* < 0·001 by Two-tailed Student’s t-test.

To simulate malnutrition conditions, we perfused the chips with ND medium which lacked the essential nutrients niacinamide and tryptophan. This resulted in villus blunting (figure 1B,C, and supplementary figure 1A), compromised barrier function (figure 1D), and reduced mucus production (figure 1E,F, and supplementary figure 1B), as previously described.^5^ Consistent with reduced barrier function, histological and immunofluorescence microscopic analysis revealed that exposure to ND resulted in epithelial and cytoskeletal damage (supplementary figure 1A), as well as partial disruption of VE cadherin-containing endothelial cell-cell contacts (supplementary figure 1C).

Considering the WHO recommendation of a high single dose (500 mg S + 25 mg P) of the SP drug combination during pregnancy, we perfused the epithelial channel of the Intestine Chip with the calculated acute duodenal dose ([133 µg S + 6·7 µg P]/mL) of this formulation as well as lower doses (1/50th [2·66 µg S + 0·13 µg P]/mL and 1/10th [13·3 S + 0·67 µg P]/mL) in culture medium and assessed its effects on morphology, barrier function, and cytokine production. We did not observe any significant changes in barrier permeability, epithelial morphology, or inflammatory cytokine production in control Intestine Chips cultured with any of the SP doses when tested in culture medium for three days (supplementary figure 2A-C). In contrast, SP treatment resulted in a significant reversal of the ND phenotype in the malnourished Intestine Chip. This was evidenced by increased villus height (figure 1B,C, and supplementary figure 1A), improved barrier function (figure 1D), enhanced mucus production (figure 1E,F), and higher levels of MUC5B (figure 1G and supplementary figure 1B). We also noted a reduction in the total number of epithelial cells in the ND condition, which was significantly reversed in the presence of SP (figure 1H).

### SP increases absorption in nutrient-deficient Intestine Chips

Nutritional components including fatty acids and vitamins help to maintain healthy gut homeostasis and prevent intestinal inflammation, and the absorption of fatty acids, zinc, and triglyceride in pregnant women is crucial for normal fetal tissue development and birth outcomes.^13,14^ When RNA-seq analysis of epithelial cells obtained from Healthy Intestine Chips, ND chips, and ND chips with SP treatment was carried out, we detected 594 genes (adjusted *p* < 0·05 and log_2_(fold change) ≥ 1·0; 149 upregulated, 445 downregulated) that were differentially regulated in the ND condition compared to healthy controls. Intestinal epithelial cells exposed to ND medium exhibited lower expression of lipoproteins (APOA1, APOC3), solute carrier (SLC) transporters (SLC2A5, SLC6A13, SLC28A1), and mucins (MUC2) compared to healthy chip controls (figure 2A). Functional enrichment analysis demonstrated that nutritional deficiency suppressed pathways related to digestion, the triglyceride metabolic process, intestinal absorption, the fatty acid metabolic process, the response to zinc ion, and the vitamin metabolic process, but activated epithelial cell fate commitment and regulation of the Wnt signaling pathway compared with healthy chip controls (figure 2B).

**Figure 2:**
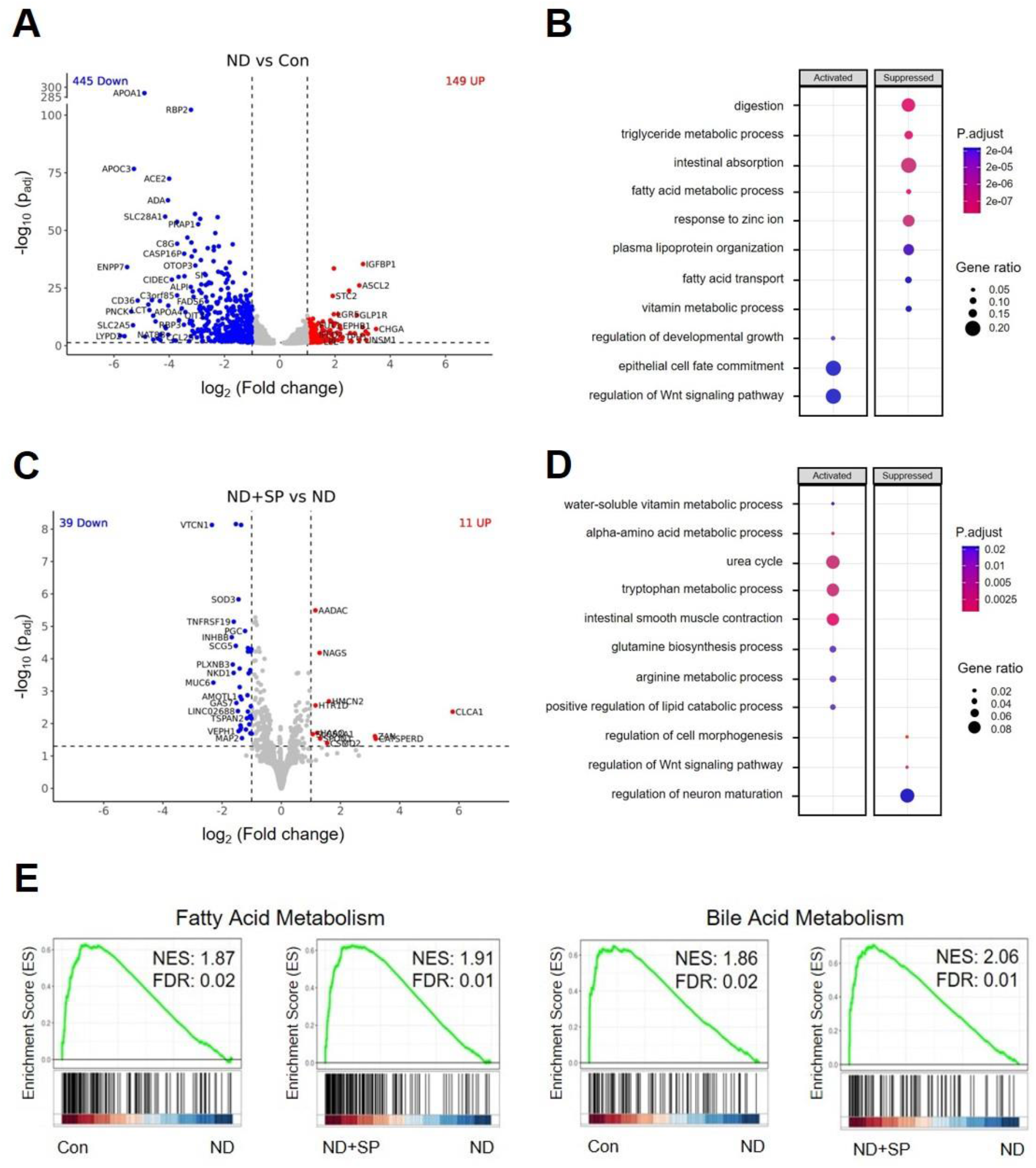
Reversal of impaired absorption in nutrient-deficient Intestine Chips by SP treatment. (A) Volcano plot of DEGs from nutritionally deficient (ND) chips compared to control (Con) chips. (B) Dot plot showing the biological processes activated or suppressed by ND vs. Con conditions from (A). (C) Volcano plot of DEGs found in epithelial cells from SP-treated ND chips (ND+SP) compared to those from ND chips. (D) Dot plot showing the biological processes activated or suppressed by ND+SP vs. ND condition from (C). (E) Gene Set Enrichment Analysis (GSEA) plots showing the significant enrichment of gene sets in epithelial cells from Con vs. ND and from ND+SP vs. ND conditions, respectively. MSigDB Hallmark 2020 was used for GSEA plot.

In contrast, when we compared transcriptomic expression profiles from SP-treated ND chips to untreated ND chips, we detected differential expression of 50 genes (adjusted *p* < 0·05 and log_2_(fold change) ≥ 1·0; 11 upregulated, 39 downregulated). The cells from ND+SP chips exhibited higher expression of various genes related to key enzymes in the small intestine (AADAC, NAGS, HMCN2), intestinal mucus homeostasis (CLCA1), a transporter for water-soluble vitamins (SLC52A1), a mediator for iron ion and oxygen binding (HAAO), those involved in cell recognition and behaviors (CATSPERD, ZAN, HTR1D), and protein processing (SPON1) during pregnancy compared to the untreated ND condition (figure 2C).^15–18^ Functional enrichment analysis indicated that cells in SP-treated ND chips reactivated various metabolic processes (water-soluble vitamin, alpha-amino acid, tryptophan, arginine, and lipid), and biological pathways (urea cycle, intestinal smooth muscle contraction, and glutamine biosynthesis), but suppressed regulation of the Wnt signaling pathway, cell morphogenesis, and neuro maturation compared with ND chips (figure 2D). Gene set enrichment analysis (GSEA) also showed that genes involved in fatty acid and bile acid metabolism are upregulated in healthy control and ND+SP chips compared to ND chips (figure 2E). Consistent with these findings, when we further analyzed cellular uptake of fatty acids using a fluorescently labeled dodecanoic acid, we found a 3·5-fold reduction in fatty acid uptake in ND chips compared to healthy chip controls, and treatment with SP significantly reversed this effect (figure 3A,B).

**Figure 3:**
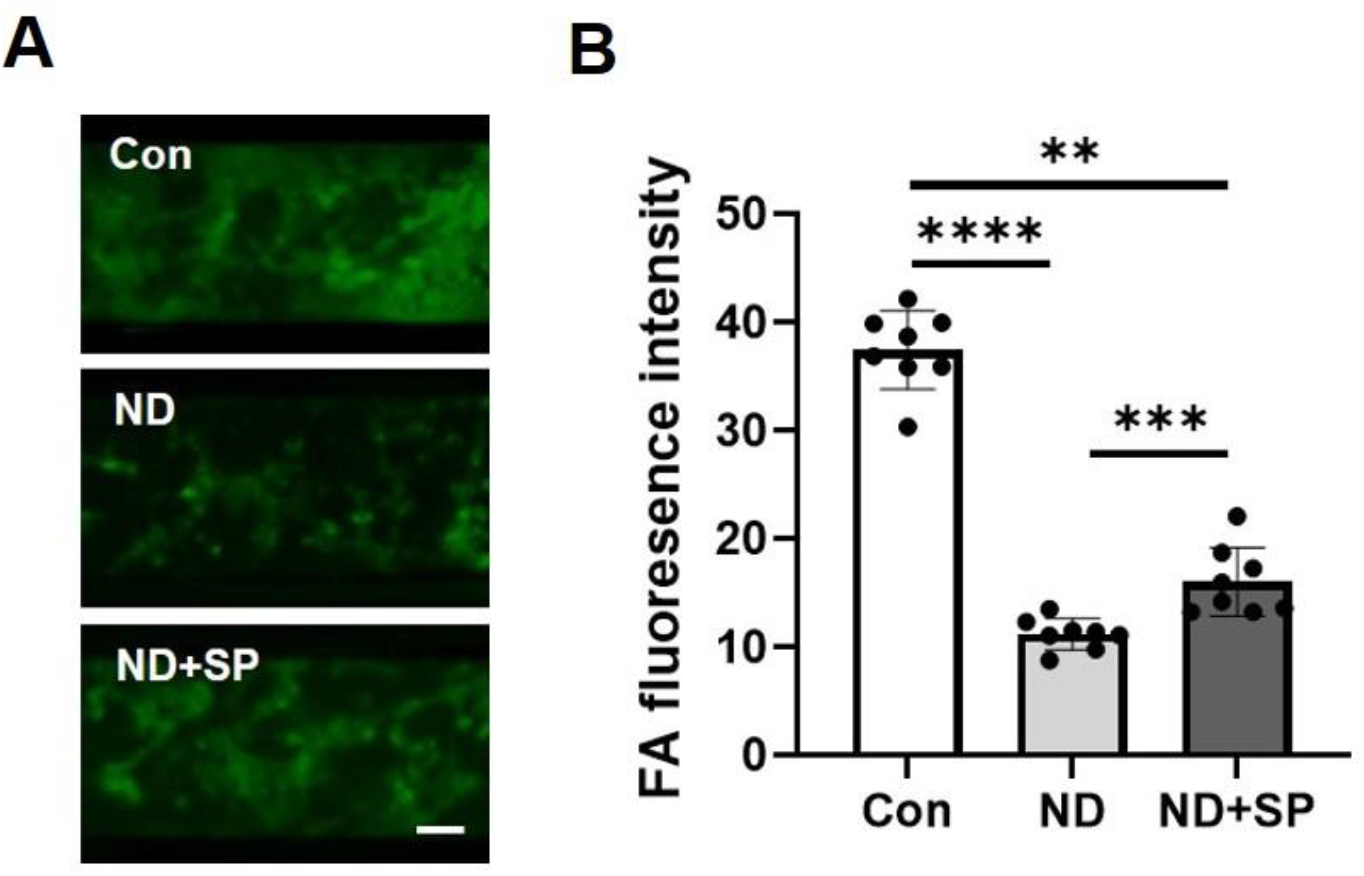
Impaired fatty acid absorption. (A) Fatty acid (FA) absorption assay in the Intestine Chips by fluorescently-labeled (Alexa 488) dodecanoic fatty acid uptake at the 60 min time point. Scale bar = 50 µm. (B) Differences in fluorescence intensity of up-taken FA. \*\**p* < 0·01, \*\*\**p* < 0·001, \*\*\*\**p* < 0·0001 by Two-tailed Student’s t-test.

### SP suppresses nutritional deficiency-associated inflammatory responses

Malnutrition-associated enteric function also commonly results in enhanced intestinal inflammation.^19^ Consistent with this clinical observation, we detected increased protein levels of multiple pro-inflammatory cytokines, including interleukin-8 (IL-8), MIP-1α, TNF-α, IFN-γ, IL-10, IL-15, REG3A, and lipocalin-2 (LCN2) in ND Intestine Chips compared to healthy control chips (figure 4A,B). Once again, treatment with SP suppressed this inflammatory response by reducing levels of most of these cytokines while further increasing the production of LCN2 (figure 4B), which has been reported to maintain intestinal microbiota homeostasis and protect against intestinal inflammation.^20^ This increase in inflammation observed in the ND chips was accompanied by epithelial damage and apoptosis, as indicated by increased Caspase-3 immunostaining, and again this was reversed by SP treatment (figure 4C,D). Furthermore, when we introduced human peripheral blood mononuclear cells (PBMCs) into the endothelium-lined vascular channel of the chip, increased numbers of these immune cells became adherent to the surface of the epithelium in ND chips compared with healthy chip controls. Importantly, this inflammatory recruitment of immune cells was also significantly inhibited by SP treatment (figure 4E,F). We also performed flow cytometric analysis of the adherent immune cells, and did not detect any significant changes in the distribution of T cells, B cells, or monocytes under the different experimental conditions (supplementary figure 3).

**Figure 4:**
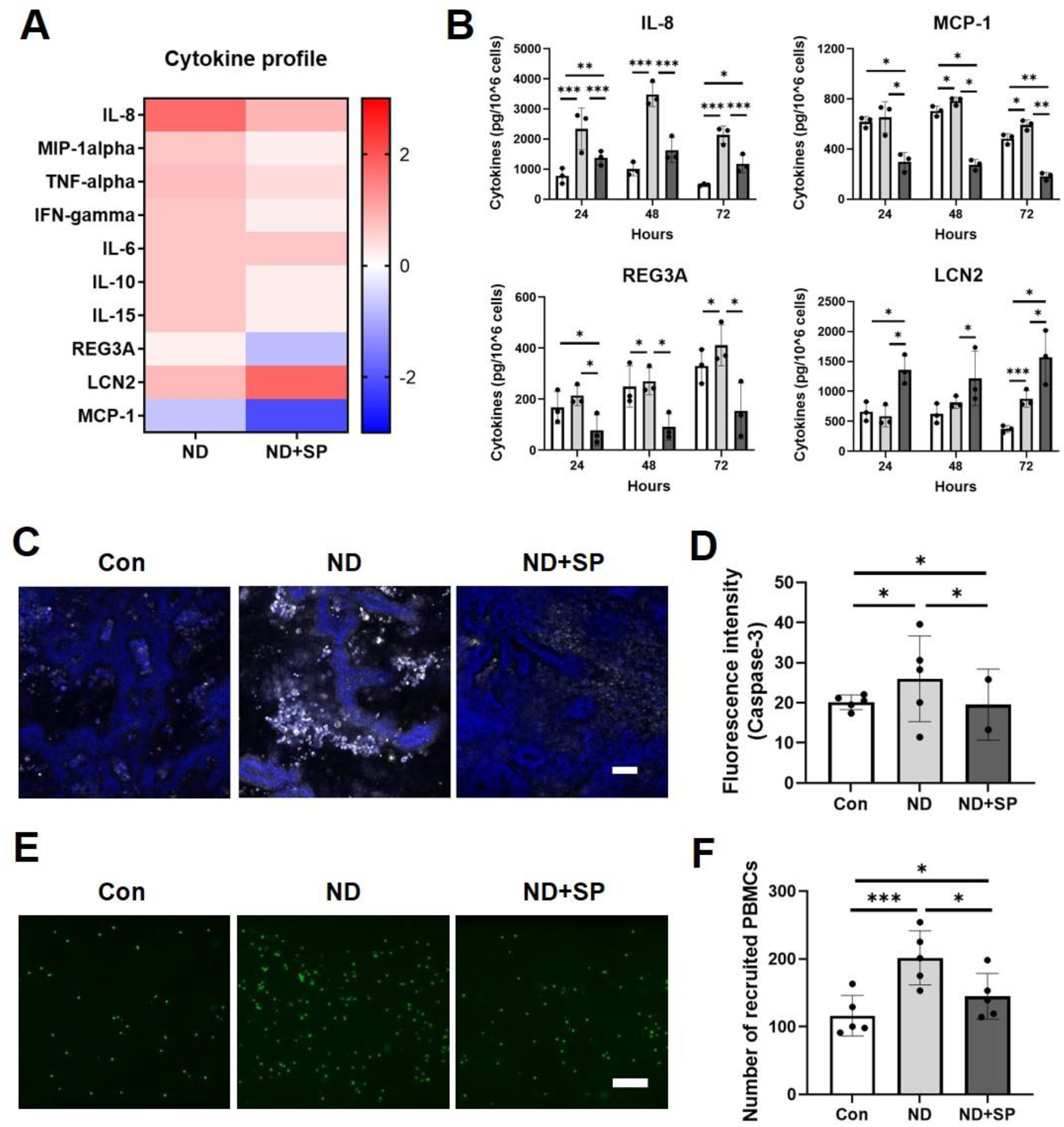
Effect of SP on inflammatory responses associated with nutritional deficiency. (A) Heatmap showing differential expression of 10 cytokines secreted into the apical (epithelium) channel of Intestine Chips at day 3 after SP treatment, measured by Luminex assay. The color-coded scale represents the log_2_ fold change in expression. (B) Production of four key cytokines on day 1, day 2, and day 3 after SP treatment. White (Con), gray (ND), and dark gray (ND+SP). \**p* < 0·05, \*\**p* < 0·01, \*\*\**p* < 0·001 by Two-tailed Student’s t-test. (C) Immunofluorescence imaging of the epithelial channel of Intestine Chips stained for Caspase-3 (white) and Hoechst 33342 (blue). Scale bar = 100 µm. (D) Differences in fluorescence intensity of Caspase-3 from Intestine Chip images (C). \**p* < 0·05, by Two-tailed Student’s t-test. (E) Recruitment of PBMCs in the Intestine Chip. PBMCs were stained with CellTracker Green^TM^ CMFDA. Scale bar = 50 µm. (F) Quantification of recruited PBMCs in the top channel of chips from Intestine Chip images (E). \**p* < 0·05, \*\*\**p* < 0·001 by Two-tailed Student’s t-test.

## DISCUSSION

The high incidence of low birth weight newborns in women with malnutrition is a critical problem in low-resource nations and a major international issue because it is associated with increased infant mortality and developmental delays in children. The serendipitous finding that SP antimalarial therapy reduces the incidence of lower birth weight infants in pregnant women in sub-Saharan Africa has raised the possibility that this antibiotic combination might have direct effects on maternal physiology that are independent of its antimicrobial actions. In the present study, we investigated this directly by leveraging Organ Chip technology to engineer human Intestine Chips lined with young adult female patient-derived duodenal epithelial cells interfaced with endothelial cells and by mimicking ND using nutrient-reduced growth medium lacking niacinamide and tryptophan. We have previously demonstrated that human Intestine Chips cultured under these ND conditions replicate the EED phenotype observed clinically.^5,21^ Importantly, our results show that SP treatment significantly improved conditions of villus blunting, compromised barrier function, mucus layer thinning, and reduced nutrient absorption, and it also prevented epithelial cell death, which are all induced by ND conditions in human Intestine Chips. SP treatment also suppresses the release of inflammatory cytokines and inhibits the recruitment of circulating immune cells. Thus, SP does indeed have direct effects on the human adult female intestine that could in part explain the improved birth weights seen in malnourished mothers who were treated with SP as an antimalarial therapy.^4,22^

SP appears to alleviate intestinal dysfunction induced by nutritional deficiencies in multiple ways based on our in vitro studies with human Intestine Chips. First, it restores a more normal villus architecture with increased intestinal surface area that is available to absorb digested food molecules. Second, SP treatment of the ND intestine also improves the secretion of mucins and increases the thickness of the intestinal mucus layer that normally lubricates the epithelial surface and protects the epithelium from commensal microorganisms, invading pathogens, and other environmental irritants.^23^ Third, SP induces the expression of multiple genes that are crucial for the efficient absorption of nutrients and fatty acids, which are extremely important during pregnancy for both maternal and fetal health.^24^ In particular, we found that ND conditions reduce fatty acid absorption in the intestinal epithelium and that treatment with SP counters this inhibition. Genes involved in fatty acid uptake, transport, and metabolism, which are important for maintaining both glucose and energy homeostasis,^25^ are also downregulated in the ND condition compared with healthy controls, and again SP treatment largely reverses this effect. Interestingly, we found that SP treatment stimulates the metabolism of bile acids, which are known to play multiple roles in lipid metabolism and serve as signaling molecules in the small intestine by modulating lipid, glucose, and energy homeostasis, in addition to supporting the absorption of dietary lipids.

In the living intestine, there is an intricate interplay between intestinal epithelial cells and the mucosal immune system,^26^ which becomes deregulated leading to increased inflammation in patients with EED or other forms of intestinal dysfunction. Similarly, we found that culturing healthy Intestine Chips lined by epithelium from young adult female donors in the presence of ND medium resulted in increased production of multiple pro-inflammatory cytokines, which is consistent with what we observed previously in chips lined by cells from EED patients when exposed to nutritional deficiency.^5^ Interestingly, after SP treatment, these elevated cytokine levels in the ND Intestine Chips reduced significantly when the chips were treated with SP. The only exception was LCN2, which was induced by SP. This is interesting because LCN2 is known to suppress inflammation in the intestine by multiple mechanisms including protecting the barrier against oxidative stress, enhancing phagocytic bacterial clearance, maintaining homeostasis with the commensal microbiome, and modulating immune responses.^27^ Consistent with these observations, we found that when we introduced PBMCs into the endothelium-lined vascular channel of the Intestine Chips, ND increased immune cell recruitment whereas SP treatment greatly reduced this response.

Taken together, these results show that SP can indeed have multiple direct effects on human intestinal structure and function that could explain the serendipitous observation of reduced incidence of low birth weight newborns, which is seen in pregnant women after anti-malaria treatment with SP. Most importantly, these findings suggest that SP should be further explored as a potential treatment for this important global health issue, and that the human Intestine Chip also may prove useful to address other questions relating to intestinal physiology and pathophysiology that are of interest to the global health community.

## METHODS

### Preparation of Intestine Chips and SP treatment

Intestinal organoids were generated from biopsy samples from healthy regions of the duodenum in patients being evaluated for potential other intestinal problems, as described,^5,8^ and all methods were performed according to the approval of the Institutional Review Board of Boston Children’s Hospital (Protocol number IRB-P00000529). Our methods for culturing patient-derived epithelial intestinal organoids, enzymatically releasing the cells, and then culturing them within one channel of a two-channel microfluidic organ chip device (Emulate Inc.) separated by a porous membrane from primary human microvascular endothelial cells in the second channel, have been described previously.^5,9^ The upper epithelial channel was perfused with complete medium or ND medium without niacinamide and tryptophan at 60 µL/h to mimic the EED state, while the lower channel was perfused with expansion culture medium. Sulfadoxine (S; 200 mg, USP) and pyrimethamine (P; 200 mg, USP) were prepared in 0·5 mL DMSO (ThermoFisher), and mixed with differentiation medium (DM) at a ratio of 20:1 (133 µg S + 6·7 µg P)/mL which simulates the acute duodenal exposure at the WHO recommended dose (500 mg S + 25 mg P/tablet) of the SP formulation (192 µg S + 9·6 µg P)/chip for 1 day; equivalent to 1 SP tablet in 250 mL gastric volume considering the gastric volume of a healthy adult (250-300 mL); these drugs were then perfused through the epithelial channel for 3 days.

### Morphological analysis

For immunofluorescence microscopic imaging, tissues within either whole or Vibratome (Leica VT1000S) sectioned chips were fixed with 4% paraformaldehyde (PFA), permeabilized, and blocked with 0.1% Triton X-100 solution and 10% donkey serum in PBS. Barrier integrity was assessed using antibodies directed against ZO-1 (Invitrogen, 33-9100), VE-Cadherin (ThermoFisher, ab33168), and species-specific secondary antibodies (Invitrogen, A31570 and Invitrogen, A31571); nuclei were stained with Hoechst 33342 (Invitrogen, H3570). Pseudo hematoxylin and eosin (H&E) staining was carried out on 40-60 µm chip sections, as described. ^10^ Imaging was carried out using a laser scanning confocal microscope (Leica SP5 X MP DMI-6000), and IMARIS (Ver. 9.9.1, Oxford Instruments) and ImageJ software were used for processing and analysis of 3D high-resolution horizontal or vertical cross-sectional images.

### Intestinal barrier permeability

We assessed the paracellular permeability of the intestinal barrier within Intestine Chips as previously described using the small fluorescent biomarker, Cascade Blue (596 Da; ThermoFisher, C687).^11^ Briefly, the apical-to-basolateral flux of the paracellular marker was calculated using the following formula: *P_app_* = (*V_r_* x *C_r_*)/ (*t* x *A* x *Cd out*), where *V_r_* is the volume of the receiving channel outflow, *C_r_* is the concentration of tracer in the receiving channel, *t* is time (sec), *A* is the total area of diffusion (cm^2^), and *Cd out* is the concentration of tracer in the dosing channel outflow (mg/mL).

### Mucus accumulation

Intestinal mucus was visualized on-chip using wheat germ agglutinin (WGA)-Alexa 488 (ThermoFisher).^5^ Briefly, 25 µg/mL of WGA in Hanks’ Balanced Salt Solution (HBSS, Gibco) was flowed at 200 μL/h through the apical channel for 30 min and then washed with continuous flow of HBSS at the same flow rate for 30 min. The entire channel was visualized with a fluorescence microscope (Excitation/Emission=488/523nm, Zeiss Axio Observer Z1). For MUC5B visualization, immunofluorescence microscopic imaging was carried out on 150-200 µm chip sections using antibodies against MUC5B (Invitrogen, 37-7400) and a secondary antibody (ThermoFisher, A31571). The MFI intensity of MUC5B staining was quantified using ImageJ.

### Fatty acid uptake

Fatty acid (FA) absorption was measured as described.^5^ Briefly, the media in the apical and basal channels of the Intestine Chips were switched to HBSS for 1 h. A fluorescently labeled dodecanoic acid containing a quencher (10 µL quencher + 5 µL FA + 485 µL assay buffer + 500 µL HBSS) was then added to the apical channel according to the manufacturer’s instructions (BioVision, K408). Fluorescence microscopic imaging of the epithelial channel was then carried out (Excitation/Emission=488/523nm, Zeiss Axio Observer Z1 and Echo Revolve microscope) and FA uptake was calculated by determining fluorescence intensity levels using ImageJ.

### Transcriptomic analysis

RNA sequencing analysis of epithelial cells removed from the chips by exposure to Accumax^TM^ (STEMCELL Technologies) was performed by AZENTA Life Sciences (NJ, USA). Briefly, RNA was isolated using RNeasy Plus Micro Kit (Qiagen) and the samples were evaluated for RNA quality. Libraries were sequenced using the Illumina HiSeq4000 platform, and the resulting150bp long pair-ended reads were trimmed using Trimmomatic (v.0.36) mapped to GRCh38 reference genome using STAR (v.2.5.2); gene hit counts to ENSEMBL Release 105 transcriptome annotation were generated using featureCounts of Subread package (v.1.5.2) for downstream analysis. Differential gene expression analysis was performed using DESeq2 (v.1.26.0) with the Benjamini and Hochberg method for adjusting *p*-values for multiple hypothesis testing. Genes with adjusted *p*-values < 0·05 and absolute log_2_(fold change) ≥ 1 were considered differentially expressed. Relevant biological pathways were identified in the MSigDB Hallmark 2020 database using gene set enrichment analysis (GSEA).^12^

### Inflammatory cytokine production

Changes in cytokine levels were measured in apical channel effluents from our chips.^11^ Ten inflammatory cytokines were selected and a Luminex assay was conducted according to the manufacturer’s protocol (Invitrogen). Analyte concentrations were measured using a Luminex FLEXMAP 3D instrument with xPONENT® software. For apoptosis analysis, cleaver Caspase-3 (Cell Signaling Technology, 9661S) was stained with a secondary antibody (ThermoFisher, A31572) and imaged with a laser scanning confocal microscope (Leica); the fluorescence intensity was measured using ImageJ.

### Immune cell recruitment

We quantified the effects of SP on the recruitment of immune cells through the endothelium and to the epithelium by perfusing PBMCs through the lower endothelium-lined channel, as previously described.^11^ Briefly, de-identified human patient-derived apheresis collars were obtained from the Crimson Biomaterials Collection Core Facility under approval obtained from the Institutional Review Board at Harvard University (#22470). Cells were stained with CellTracker Green^TM^ CMFDA (1:1,000 v/v in PBS, ThermoFisher, #C7025) for 10 min at 37°C in a water bath and stained PBMCs were then seeded into the basal channel (endothelium) of the Intestine Chips at 5 × 10^7^ cells/mL in medium. PBMCs were allowed to adhere to the endothelium by inverting the chips for 3 h under a static condition, were gently washed with complete medium to remove non-adherent or suspended cells, and were then maintained at continuous flow (60 µL/h). The number of PBMCs that adhered and migrated from the basal vascular channel to the apical epithelium-lined channel was quantified by immunofluorescence microscopic imaging. The composition of these PBMCs recruited to the epithelium was determined using Flow Cytometry (CytoFlex LX); cell digests were stained with Viakrome 808 Live/Dead stain (Beckman Coulter, C36628), CD19 BV421 (BioLegend, 302234), CD3 APC-H7(BD Biosciences, 560176), and CD14 BV395 (BD Biosciences, 563561), and then fixed with 200 µL of Cytofix (BD Biosciences, 554655). To calculate the absolute number of cells per sample, 5025 counting beads were added to each sample. Results were analyzed using FlowJo V10 software (FlowJo, LLC).

### Statistical analysis

Between 3 and 6 Intestine Chips were used in each study and a Two-tailed Student’s t-test was performed to determine statistical significance, as indicated in the figure legends. *P <* 0·05 were considered significant; error bars indicate mean ± standard deviation (s.d.).

## Data Availability

The datasets and analysis will be available upon request. All requests should be addressed to the corresponding author.

## Acknowledgments

This study was funded by the Bill and Melinda Gates Foundation and the Wyss Institute for Biologically Inspired Engineering at Harvard University. We would like to thank Thomas C. Ferrante and Alican Ozkan for their help in processing the images. We also thank Gwenn E. Merry for editing this manuscript.

## Contributors

S.K. led this study, and S.K., G.G., and D.E.I. designed the overall research. S.K., A.N., and P.P. performed experiments. S.K., V.H., A.J., G.G., and D.E.I. analyzed and interpreted all the data. D.T.B. established human intestinal organoids. S.K. and D.E.I. wrote the article with input from other authors.

## Supplementary Data

**Supplementary Figure 1:**
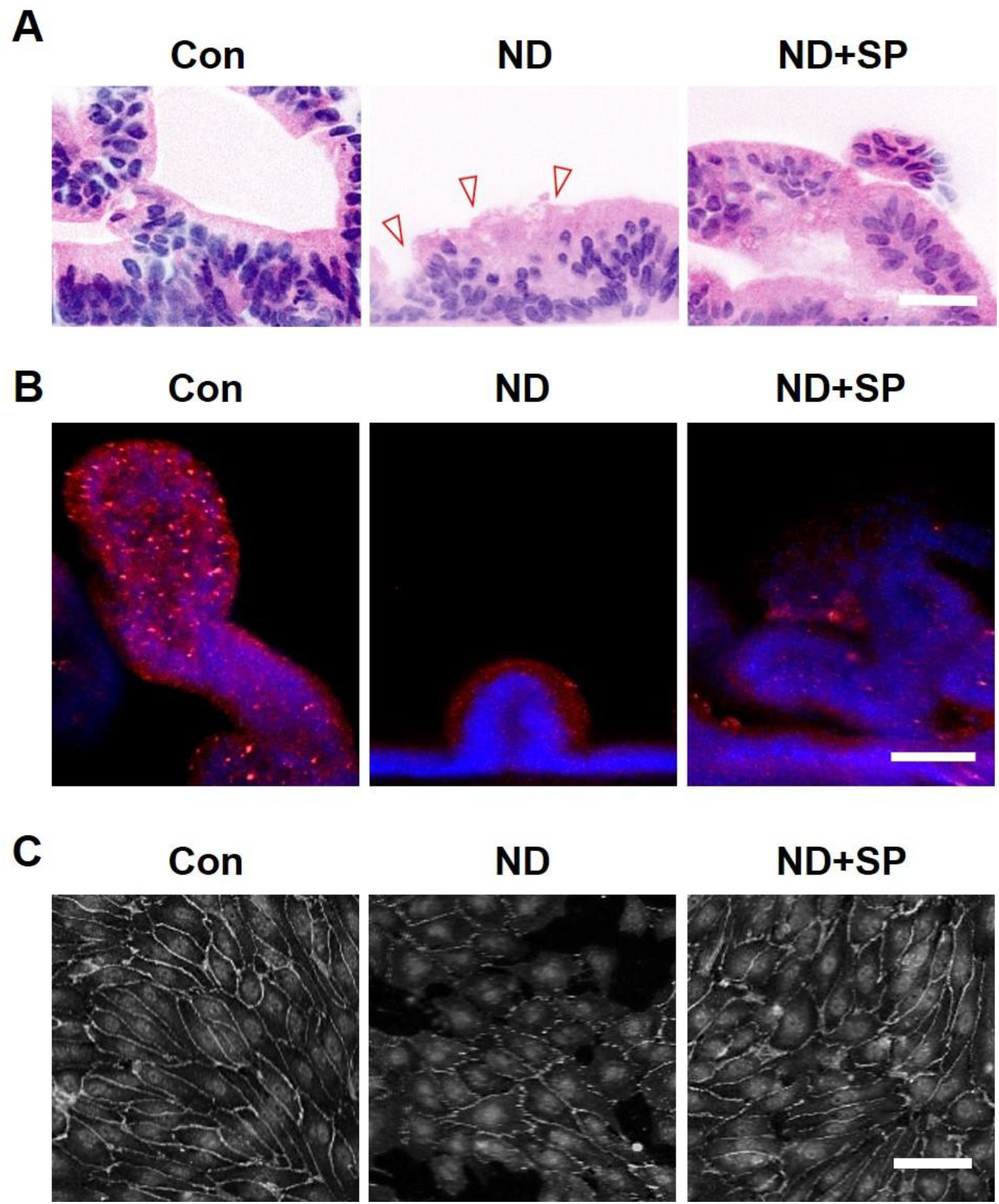
Morphological characteristics of Intestine Chips. (A) Pseudo-H&E stained images of the intestinal epithelium on Intestine Chips. Red arrows indicate regions of potentially damaged epithelium and pseudostratified cytoskeleton. Scale bar = 50 µm. (B) Immunofluorescence imaging of cross-sectioned Intestine Chips showing the presence of mucin, red: MUC5B, blue: Hoechst 33342. Scale bar = 50 µm. The fluorescence intensity of MUC5B is shown in figure 1G. (C) Immunofluorescence imaging of the basal channel (endothelium) of Intestine Chips stained with VE-Cadherin (white) and Hoechst (gray). Scale bar = 50 µm.

**Supplementary Figure 2:**
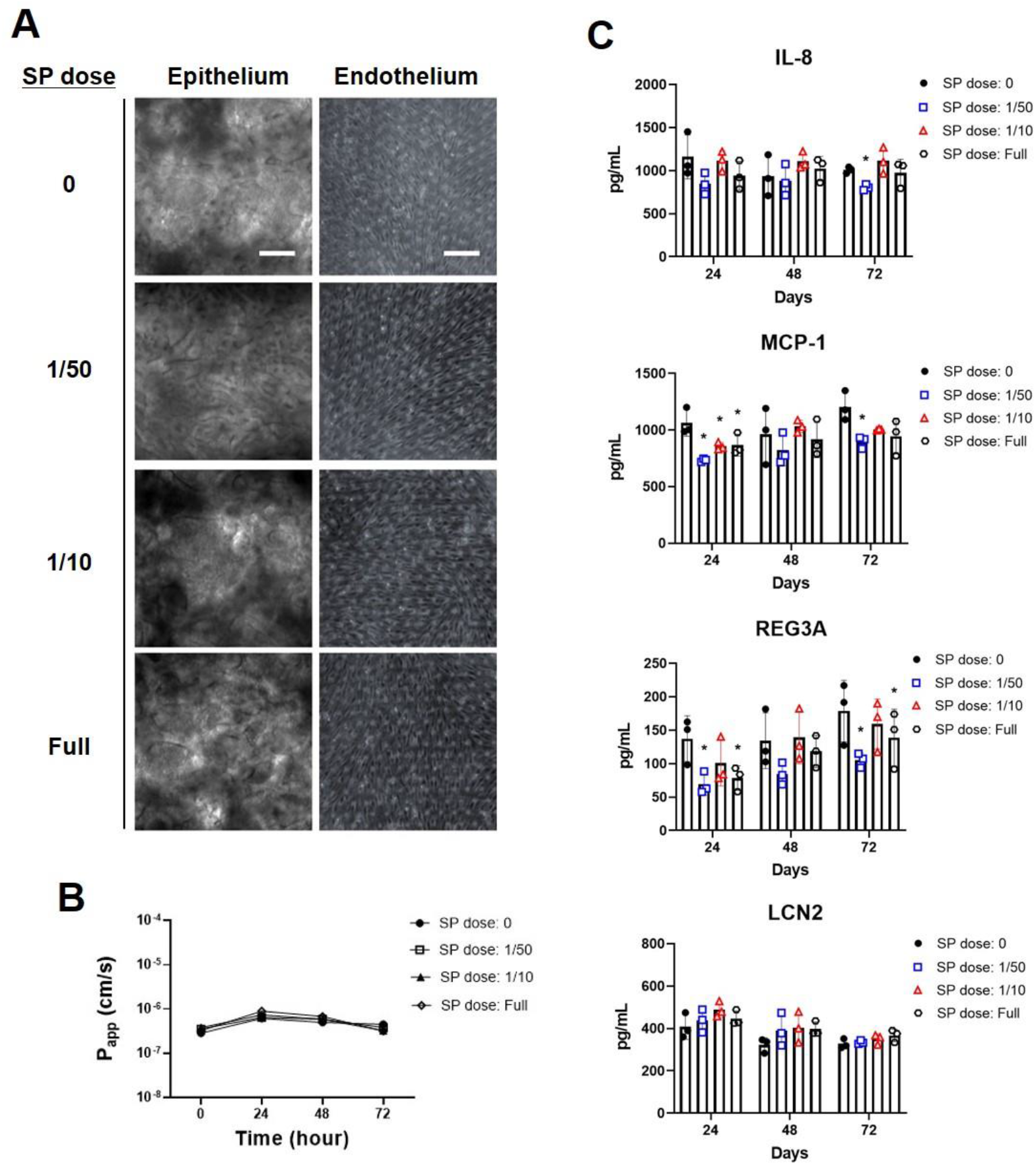
Optimal dose-finding for an Intestine Chip model. (A) Phase-contrast imaging of Intestine Chips. Intestine Chips were treated with DMSO only (control) or different doses of SP dissolved in DMSO. The epithelial channel of the Intestine Chip was perfused with the calculated acute duodenal dose ([133 µg S + 6·7 µg P]/mL) of this formulation as well as lower doses (1/50th [2·66 µg S + 0·13 µg P]/mL and 1/10th [13·3 S + 0·67 µg P]/mL) in culture medium. Scale bar = 100 µm. (B) Apparent permeability (*P_app_*) after SP treatment with different doses. (C) Cytokine analysis at day 1, day 2, and day 3 after SP treatment was measured by Luminex assay in the effluent from the apical (epithelium) channel of Intestine Chips. \**p* < 0·05 by Two-tailed Student’s t-test.

**Supplementary Figure 3:**
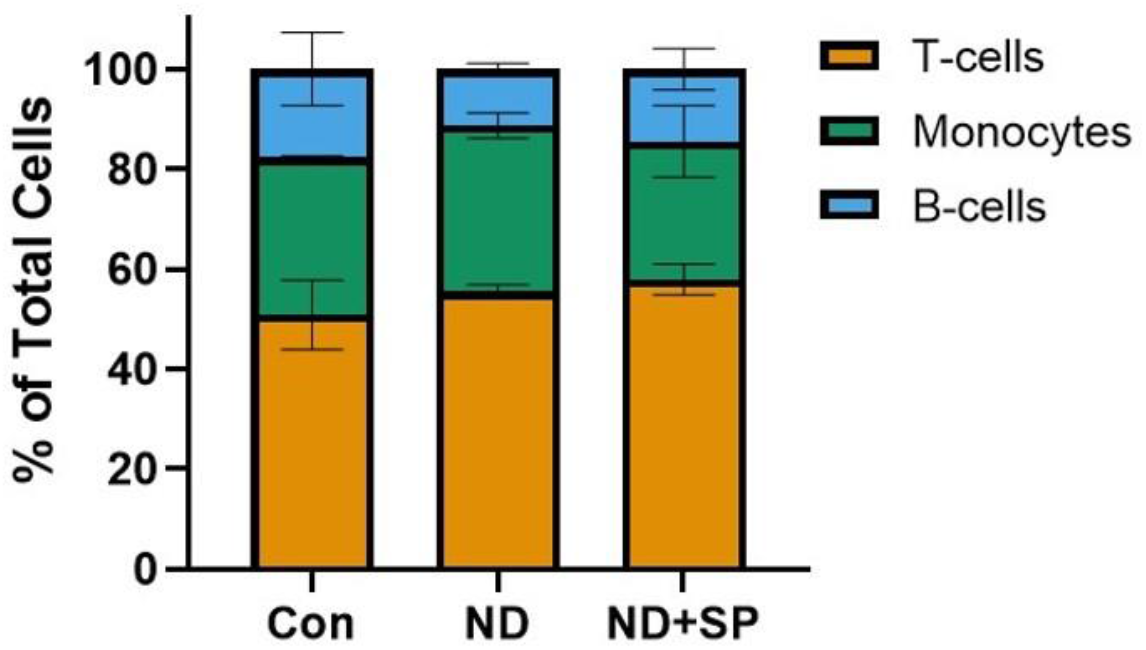
Composition of recruited immune cells. PBMCs were collected from Intestine Chips and identified by FACS. Orange (T-cells by CD3), green (Monocytes by CD14), and blue (B-cells by CD19).

